# Did State-level telehealth policies in 2020 Reduce Urban-rural Disparity in Care Utilization? A Multilevel Analysis

**DOI:** 10.64898/2026.02.08.26345818

**Authors:** Lu Shi, Yucheng Wang, Corey J. Hayes, Cari A. Bogulski, KaSheena Winston, Denise Tahara, Hari Eswaran

## Abstract

**Introduction:** We aim to examine whether state-level telehealth policies in 2020 were associated with a reduction in the urban-rural disparity in telehealth utilization.

**Method:** A multilevel model was used to assess state actions’ impact on urban-rural differences in healthcare utilization. We used the percentage of Medicare Fee-for-Service beneficiaries receiving any telehealth services (measured by county level) in 2020 as the dependent variable. We examined the following state-level policies as key independent variables at the state level: 1) telehealth coverage parity requirement for payers; 2) recognizing a call from patient’s address as the originating site of telehealth visit; 3) mandating the coverage of audio-only telehealth visits.

**Results:** Both mandating the coverage of audio-only visits and telehealth coverage parity requirement were significantly associated with higher level of telehealth utilization. For audio-only reimbursement mandate and telehealth coverage parity requirement, the interaction between the state policy and the county’s rurality was associated with a significant increase in telehealth utilization rate, whereas the interaction between the state’s waiver for the requirement of originating site and the county’s rurality was negatively associated with telehealth utilization rate.

**Conclusion:** The audio-only telecare reimbursement mandate and telehealth payment parity could help close the urban-rural gap in telehealth utilization.

## Introduction

In 2020 when the Covid-19 Pandemic threatened the safety of in-person outpatient visits, many state governments issued a series of emergency policies to facilitate telehealth utilization. Two of the most widely adopted policies during this time were telehealth coverage mandates for private payers^1^ and emergency licensure waivers to allow out-of-state providers to have a healthcare visit with patients in a different state^2^. Other policy initiatives include allowing Federally Qualified Health Centers (FQHCs) and Rural Health Centers (RHCs) to provide telehealth services^3^, requiring telehealth parity for payers^4^, recognizing a call from a patient’s address as the originating site as a telehealth visit^5^, covering audio-only telehealth visits^6^, and suspending the limits for tele-supervised clinical practice by junior clinicians^7^. At the federal level, the Center for Medicare and Medicaid Services also issued temporary flexibilities such as allowing reimbursement of audio-only telehealth services^8^ while Federal Communications Commission (FCC) initiated extensive efforts to enhance access to telehealth services^9^.

It is plausible, then, that the urban-rural disparity in telehealth utilization researchers observed in the initial phase of the Covid-19 pandemic^10^ might have been reduced by these emergency investments, waivers and flexibilities, as policies such as covering audio-only telehealth visits could help rural patients where telecommunication signals might be too weak for audio-visual telehealth visits. However, not all states issued telehealth coverage mandates and licensure waivers, while the duration of these mandates and waivers also varied between states. For instance, only one third of the states issued mandates for covering audio-only telehealth services, one third of the states required reimbursement parity for telehealth service and two thirds of the states required recognizing the patient address as an originating site^11^. These between-state variations in telehealth policy provide us with a unique opportunity to examine whether these telehealth policy initiatives at the state level would attenuate the urban-rural disparity in healthcare utilization during the Covid-19 pandemic, an issue very relevant for future policy regarding telehealth regulation beyond the Covid-19 pandemic.

As rural America was found to be particularly challenged with utilizing the telehealth services in 2020^12^, it is important to identify potential enabling factors that were associated with alleviation of this urban-rural disparity pattern at a time of the need for telehealth visits were urgent. The need to understand these possible enabling factors becomes particularly relevant when telehealth interventions in rural communities have been shown as effective in improving patient satisfaction as well as saving patients travel time and cost^13^. As for health outcomes, recent evidence has shown the noninferiority of audio-only telehealth services as compared with video telehealth services in disease management of diabetes, hypertension and renal disease^14^, meaning that maintaining audio-only telehealth service for these patient populations could be a particularly cost-effective strategy as compared with more resource-intensive video telehealth. In this study, we examine the potential role of state-level telehealth policies in reducing urban-rural disparity in healthcare utilization during the Covid-19 pandemic from January 2020 through September 2020. We hypothesized urban-rural disparities would be lower in states that issued the following three policies in 2020: coverage of audio-only telehealth visits, origination site being a patient’s home, and telehealth coverage parity requirement for payers.

## Method

### Outcome measure: county-level telehealth utilization rate

We used the percentage of Medicare Fee-for-Service beneficiaries who received any telehealth services (measured at the county level) from January 2020 through September 2020. These county-level rates of telehealth utilization were obtained from the published estimates by CareJourney (using claims data)^15,16^, a healthcare analytics company that conducts market assessments and identifies target populations in order to improve of patient outcomes. While those state-level policies we examined in this study did not directly influence reimbursement practice for the Medicare patients covered by our sample, it is plausible that Medicare beneficiaries could still benefit from these state-level initiatives through their local providers’ telehealth service expansion and telehealth training enhancement in response to those insurance policies^17^. The geospatial spillover effect of healthcare innovation has been observed among healthcare providers serving Medicare patients in the United States^18^.

### Research Questions

We examine the effectiveness of the following state-level telehealth policies in reducing urban-rural disparity in telehealth utilization:

1. Telehealth coverage parity requirement for payers, whereby payers must offer the same level of coverage for telehealth services as they do for in-person services^4,19^
2. Recognizing a call from a patient’s address as the originating site as a reimbursable telehealth visit^5^
3. Cover audio-only telehealth visits as a reimbursable healthcare utilization^6^

We use a multilevel model to assess state actions’ impact on urban-rural difference in healthcare utilization, using the state level as the cluster level. The key independent variables will be the following three binary state-level policy variables, each representing a state-level policy decision:

1. Whether the state had a state-level mandate on telehealth reimbursement parity in 2020
2. Whether the state had an action to include audio-only phone in 2020
3. Whether the state required the payer to recognize the patient address as originating sites of telehealth visit in 2020

These state-level policy variables were coded based upon the Brookins Institution’s compilation of state actions on telehealth during the Covid-19 pandemic^11^.

Our multilevel model uses state-level policy variables and county-level covariates to control for the confounding factors and uses the interaction terms of state policy variables and county-level rurality (percent rural residents in the county^20^) to measure the interaction between these state-level policies and a county’s rurality on the telehealth utilization among Medicare beneficiaries. A positive and statistically significant coefficient for an interaction term would indicate that this policy was associated with more increase among Medicare beneficiaries in more rural counties. The county-level covariates were selected from the county-level variables compiled by University of Wisconsin Madison’s County Health Ranking^21^, using the least-angle regressor selector function in SAS 9.4’s Proc GLMSELECT module. The Proc Mixed procedure of SAS 9.4^22^ was used for our multi-level analysis.

## Results

A total of 3,202 counties nationwide were included in our analysis. Table 1 charts the descriptive statistics of the variables we used for the analysis.

**Table 1:** Descriptive statistics of state and county-level variables.

| Variables | Mean (Std) for continuous variables or % for categorical variables |
| --- | --- |
| N, State | 50 |
| Characteristics of states |  |
| Requiring Insurers Cover Audio-Only |  |
| Yes | 35.56 |
| No | 64.44 |
| Requirement for parity in provider reimbursement |  |
| Yes | 66.55 |
| No | 33.45 |
| Recognizing a call from patient address as the originating site |  |
| Yes | 32.27 |
| No | 67.73 |
| % Rural Population at the state level | 28.36 (12.03) |
| N, Counties | 3126 |
| Characteristics of counties |  |
| Telehealth utilization rate (%) | 22.22 (8.65) |
| Resident population | 206379 (1263492) |
| % of less than 18 years of age | 21.83 (3.48) |
| % 65 and over | 20.18 (4.87) |
| % Black | 9.06 (14.22) |
| % American Indian and Alaska Native | 2.41 (7.72) |
| % Asian | 1.67 (3.09) |
| % Hispanic | 9.97 (13.87) |
| % Non-Hispanic White | 75.41 (20.22) |
| % Female | 49.91 (2.22) |
| % population living in a rural area | 58.10 (31.53) |
| Number of deaths before 75y | 2825.7 (15327.3) |
| Years of Potential Life Lost Rate per 100,000 | 8970.9 (2895.2) |
| % Fair or Poor Health | 20.57 (5.02) |
| Number of Physical Unhealthy Days in 30 days | 4.33 (0.75) |
| Num Mental Unhealthy Days in past 30 days | 4.88 (0.69) |
| % Low Birthweight | 8.21 (2.03) |
| % Smokers | 20.31 (4.21) |
| % Adults with Obesity | 35.67 (4.33) |
| % with access to physical activity place | 55.45 (23.80) |
| # of motor vehicle deaths | 118 (698.5) |
| % of driving deaths with alcohol | 27.51 (15.05) |
| Discharges for Ambulatory Care with Sensitive Conditions per 100,000 Medicare Enrollees | 4037.4 (1542.6) |
| % Vaccinated | 43.00 (10.02) |
| % Unemployed | 6.75 (2.28) |
| Log Percentile 80th Income | 11.54 (0.21) |
| Social Association Rate | 11.43 (5.86) |
| Severe Housing Cost Burden | 10.56 (3.46) |
| % Long Commute Drives Alone | 32.37 (12.69) |

Table 2 presents the multilevel model we run with SAS Proc mixed. While a state’s overall rurality (measured by percent rural at the state level) was not significantly associated with county-level telehealth utilization rate among the Medicare fee-for-service population, the county-level rurality was negatively associated with this county-level telehealth utilization rate with very strong statistical significance (β= −0.06, p<.0001, 95% Confidence Interval: −0.08, - 0.04). At the state level, both mandating the coverage of audio-only visits and telehealth coverage parity requirement were significantly associated with higher level of telehealth utilization among the state’s counties (Audio-only coverage mandate: β=15.7, 95% CI=6.42, 81.00; Telehealth reimbursement parity requirement: β=9.29, 95% CI=3.79, 47.60). The state-level waiver for originating site requirement was not significantly associated with county-level telehealth utilization rate.

**Table 2:** Multilevel model of County-level Telehealth Utilization Rate among Medicare Fee-for-service Beneficiaries in 2020.

| **County-level parameters** | **$\beta$** | **P** | **Lower** | **Upper** |
| --- | --- | --- | --- | --- |
| % Hispanic | 0.08 | <.0001 | 0.06 | 0.11 |
| % Non-Hispanic White | 0.04 | <.0001 | 0.02 | 0.06 |
| % living in a rural area | -0.06 | <.0001 | -0.08 | -0.04 |
| % with access to physical activity place | 0.03 | 0.00 | 0.01 | 0.05 |
| # of motor vehicle deaths | 0.00 | 0.92 | 0.00 | 0.00 |
| % of driving deaths with alcohol | -0.01 | 0.43 | -0.03 | 0.01 |
| Discharges for Ambulatory Care with Sensitive Conditions per 100,000 Medicare Enrollees | 0.00 | <.0001 | 0.00 | 0.00 |
| % Medicare enrollees with annual flu vaccination | 0.21 | <.0001 | 0.17 | 0.25 |
| % of population ages 16+ unemployed and looking for work | 0.17 | 0.02 | 0.02 | 0.31 |
| Log 80th percentile of median household income | 10.21 | <.0001 | 8.62 | 11.79 |
| Associations per 10,000 population | -0.29 | <.0001 | -0.36 | -0.21 |
| % of households with high housing costs | 0.24 | <.0001 | 0.15 | 0.32 |
| % commuting more than 30 minutes among workers driving alone | 0.07 | <.0001 | 0.05 | 0.09 |
| **State-level parameters effects** |  |  |  |  |
| Audio-only coverage mandate | 15.75 | <0.001 | 6.42 | 81.00 |
| Telehealth reimbursement parity | 9.29 | <0.001 | 3.79 | 47.60 |
| Waiver for originating site requirement | 8.36 | 0.20 | -1.92 | 1415.46 |
| % Rural population in the state | 0.01 | 0.08 | 0.00 | 0.05 |
| **Interaction terms between state policy variables and county' rurality** |  |  |  |  |
| Audio-only coverage mandate* % Rural | 0.03 | 0.00 | 0.01 | 0.05 |
| Telehealth reimbursement parity* % Rural | 0.02 | 0.05 | 0.00 | 0.03 |
| Waiver for originating site requirement * % Rural | -0.02 | 0.01 | -0.04 | -0.01 |

The interaction between mandating the reimbursement of audio-only telehealth visits and the county’s rurality was associated with a statistically significant increase in telehealth utilization rate in the county (β =0.03, 95%CI: 0.010, 0.050). The interaction between the state’s payment parity and the county’s rurality was positively associated with the county’s telehealth utilization rate among the Medicare fee-for-service population (β =0.02, 95%CI: 0.004, 0.030). whereas the interaction between the state’s waiver for the requirement of originating site and the county’s rurality was negatively associated with telehealth utilization rate at the county level (β = −0.02, 95% CI: −0.04, −0.01).

## Discussion

Our study found that requiring the insurers to pay for audio-only telehealth services, a policy that aims to facilitate telehealth adoption among those who are not yet ready for tele-video health services, might have helped rural counties more than urban counties in improving telehealth utilization rate among Medicare fee-for-service populations, thus potentially reducing the urban-rural disparity in telehealth utilization for a patient population who were at elevated risk of infectious diseases associated with in-person visits in 2020. Similarly, the interaction between the reimbursement parity rule for telehealth services at the state level and the county’s rurality is also found to be positively associated with telehealth utilization rate among Medicare fee-for-service populations (in addition to this parity rule’s significant main effect on telehealth utilization), signaling that the reimbursement parity rule for telehealth service could also help reduce the urban-rural gap in telehealth utilization. It is important to note that these state-level payer mandates did not directly affect the payment for Medicare fee-for-service programs, yet it is plausible that many of the Medicare fee-for-service beneficiaries could nevertheless benefit from those state-level actions which could help providers adopt more telehealth services^17^. As the healthcare stakeholders consider and debate about whether pandemic-time emergency regulations and deregulations need to be terminated, it is important to weigh in these policies’ role in reducing urban-rural disparity in telehealth utilization and whether terminating some of these policies could increase the urban-rural disparity, given that telehealth could play a strong role in serving the needs of rural populations.

Our findings about audio-only telehealth’s role in rural areas is consistent with prior empirical evidence that eliminating audio-only telehealth service would disproportionately affect patient population already at disadvantage for health care access^23^. Our finding about telehealth reimbursement parity law’s significant role in elevating telehealth utilization, both as a main effect for all and as an interaction effect for rural patients, contributes to the emerging evidence base that telehealth reimbursement parity law is associated with healthcare utilization improvement (e.g., better antihypertensive medication adherence^24^) at the population level.

On the other hand, our multilevel model also shows that lifting the restriction for telehealth originating site, another state-level emergency response policy in 2020, seemed to help the urban counties more than those rural counties. Until 2019, Medicare covered telehealth services only when they were provided from an originating site outside a Metropolitan Statistical Area or in a rural Health Professional Shortage Area (HPSA) in a rural census tract^25^. During the COVID-19 pandemic, the qualifications of an eligible originating site, such as eligibility of a service received by the patient while at home, were expanded^26,27^. Because the rurality pre-condition for telehealth was removed to expand originating sites beyond rural facilities, it is perhaps unsurprising that originating site policy changes increased the uptake of telehealth services more in counties with higher urban than rural populations, especially in therapeutic areas where telehealth or telehealth-assisted home healthcare might have an advantage over in-person care ^25^. The fact that urban counties seemed to benefit more from the eligibility expansion for telehealth originating site can also be interpreted as a possible result of existing urban-rural gap in telehealth adoption and infrastructure among healthcare providers and patients^28,29^, and therefore further investment in telehealth services among rural providers could be useful in reducing the urban-rural gap in telehealth utilization.

To the best of our knowledge, our proposed study is among the earliest attempts to understand the emergency legal and executive actions’ impact on reducing urban-rural disparity in telehealth. Our findings from the Medicare population bear relevance for the elderly and disabled population’s access to healthcare: for those who might have impairment that hinders their driving capability, the telehealth option might be particularly useful for rural residents due to the limited availability of public transportation and taxi service in rural areas.

Worldwide, rapid adoption of telehealth by health providers due to lockdowns and social distancing has been noted during the COVID-19 pandemic^30^ and telehealth has become more of a critical mechanism to deliver care. However, “the digital divide” between those who have access to technology and those who do not^30^ persisted between urban and rural areas, with structural barriers like inadequate telecommunication infrastructure and individual-level barriers like inadequate digital competency. Minorities, non-English speakers, seniors and Medicaid beneficiaries are least likely to have sufficient digital literacy to engage in video telehealth options^31-33^.

While seniors, persons with disabilities, and those who are frail or have mobility challenges would especially benefit from telehealth visits, they need more confidence and assistance in engaging with the technology^31^. Covering audio-only telehealth visits therefore enabled healthcare access for many at-risk populations who, without this option, would not utilize much-needed healthcare services. The hardware and digital competency expected of video visits will be more challenging for some of these at-risk populations. To increase use of video visits, practitioners will need to provide assistance and support to increase patients’ confidence and ability to engage through telehealth visits^31^. These strategies combined with increased investment to mitigate structural barriers, particularly in rural communities, would improve telehealth utilization, patient engagement, and health equity.

Our study is limited in that our sample was drawn from the Medicare Fee-for-service patient population, who were known to have a healthcare utilization pattern different from other segment of Medicare beneficiaries (e.g., Medicare Advantage patients)^34^ and those not covered by Medicare. Therefore, our finding cannot be directly generalized to other populations with different insurance status. Considering that telehealth delivery models could have their unique temporal pattern of expenses (e.g., high set-up costs yet cost-effective in the long run)^35^, their feasibility could vary across different payment plans and therefore separate analyses will be needed for population with commercial insurance, Medicaid, Medicare population with managed care plans as well as those uninsured.

Our study did not consider the status of Interstate Medical Licensing Compacts^36^ as a pandemic-time state telehealth policy and thus did not include them in the analysis. These compacts include Psychology Interjurisdictional Compact (PSYPACT)^37^, Nurse Licensure Compact (NLC)^38^, Physical Therapy Licensure Compact (PTLC)^39^, Recognition of EMS Personnel Licensure Interstate Compact (REPLICA)^40^, and Audiology and Speech-Language Pathology Interstate Compact ^41^. While these compacts could have facilitated telehealth practice across the state borders, many of these compacts had existed long before the pandemic and the new adopters might not need to immediately join the compacts at the early phase of the Covid-19 pandemic in the same manner as state legislatures passed emergency measures as a rapid response to the ongoing pandemic. As such, the impact of these interstate compacts on telehealth utilization could not easily be assessed with our sample, which only included the utilization measure in the calendar year of 2020. Furthermore, the actual impact of these interstate medical licensing compacts on care utilization could vary based on the spatial location of the counties: the care utilization behavior could be affected more in a border county than in a non-border county, and counties in smaller states such as Delaware might be affected more than those in larger states such as Texas. Therefore, we will need geospatial models that address these geospatial patterns to better understand the actual impact of these interstate medical licensing compacts.

The Covid-19 pandemic-epidemic, now lasting in into its sixth calendar year (since the initial human infection documented in 2019), has led to notable growth in telehealth services around the world. National governments and local authorities have taken a series of emergency legislative and executive moves to facilitate the telehealth service for the purpose of minimizing infection during healthcare visits while maintaining healthcare during the pandemic time. With many of these state actions likely to expire as the vaccination campaign finally decreased the infection risk, for the purpose of reducing urban-rural disparity in telehealth utilization it is important to consider which one of these emergency actions played a role in reducing the urban-rural disparity in telehealth utilization, as stakeholders weigh the decisions of whether to make some of these policies permanent in the post-pandemic era.

## Data Availability

All data produced in the present study are available upon reasonable request to the authors

